# Caregiving burden and associated factors among family caregivers of individuals with schizophrenia in Bangladesh: A cross-sectional study

**DOI:** 10.1101/2023.06.01.23290855

**Authors:** Tahsin Tasneem Tabassum, Nur-A-Safrina Rahman, S M Shakhawat Hossain, Faisal Abdullah, Lubaba Tasneem Nawar, Fahmida Islam Lima, Mridul Gupta, Suraiya Parveen Kona, Vivek Podder

## Abstract

**Background:** Schizophrenia is a severe mental disorder affecting millions worldwide. Family caregivers play a vital role in patient treatment and recovery, but their responsibilities can be physically and emotionally exhausting. There is limited research on caregiver burden in Bangladesh.

**Purpose:** This study to identify factors associated with the burden among caregivers of schizophrenic individuals.

**Methods:** This cross-sectional study collected data from 175 caregivers from January to December 2020 in Dhaka, Bangladesh. A semi-structured questionnaire and validated Bangla version of the Zarit Interview Burden scale were used to assess the burden of primary caregivers and associated factors in caring for patients with schizophrenia.

**Results:** Participants’ mean age was 34.02±10.45 years, with 26.9% in the 34-41 age group. 49.1% were housewives, with most earning 5000 to 15000 taka monthly. Spouses accounted for 28.6% of caregivers. Most patients had an illness duration of less than five years (66.3%). Mean ZBI score was 49.49±12.06, indicating moderate to severe burden. Factors significantly associated with caregiver burden included age, gender, occupation, income, marital status, house condition, relationship with patients, illness stage, and duration.

**Conclusion:** This study highlights the burden experienced by caregivers of schizophrenic patients in Bangladesh and identifies factors associated with the burden. It recommends community interventions and psychosocial provisions to address this issue and inform targeted interventions to reduce caregiver burden. These findings provide insights for a comprehensive plan to manage such cases in the future.

## Introduction

Schizophrenia is a chronic, severe mental disorder that affects millions of people worldwide, with a prevalence rate of approximately 1 in 300 people (0.32%).^1^ Among adults, this rate is even higher at 1 in 222 people (0.45%).^1^ Schizophrenia has a high prevalence rate of 1.1% in adults, with an estimated 51 million people worldwide suffering from the disorder.^1^ The prevalence of schizophrenia in Bangladesh is 1.10% among adults and 0.10% among children.^2^ A study conducted in a rural community found the prevalence rate of schizophrenia to be 2.54 per 1000 of the population.^3^ The condition can cause significant impairment in individuals’ ability to function, including their social and occupational functioning, and has a significant impact on the lives of their family members, particularly primary caregivers. It is a chronic condition that requires ongoing management, which can be challenging for both patients and their caregivers. Caregivers of individuals with schizophrenia play a vital role in their treatment and recovery, but this responsibility can also be emotionally and physically exhausting. After the deinstitutionalization policies, the burden of taking care of psychiatric patients fell largely on their families. In Iran, between 65% and 75% of discharged schizophrenia patients return to their families for care.^4^ Research shows that these patients often require 24-hour care and cannot stay alone for more than 3 hours.^5, 6^ Schizophrenia presents specific caregiving needs, and family caregivers have multiple roles in caring for patients, including assisting with daily and self-care activities and managing unexpected behaviors such as aggression and violence. The responsibilities imposed on family caregivers can result in a significant burden, with 83% to 95% of caregivers experiencing decreased quality of life and increased risks for both caregivers and patients.^7^

Families of individuals with schizophrenia have been studied for decades. As care shifted from hospitals to homes, families took on a major portion of caregiving responsibilities, leading to the concept of “burden of care.”^8^ Caregiver burden is the emotional, physical, and financial demands placed on family members, friends, or others involved in caring for the individual.^8, 9^ Long-term caregiving can lead to stress, decreased quality of life, and lack of support from mental healthcare providers.^10, 11^ Many studies have been conducted to determine factors associated with the burden of primary caregivers of patients with schizophrenia.^12, 13, 14^ The number of patient needs, higher levels of psychopathology and disability, being male and older, patients’ symptoms, male gender, unemployment and marital status, and caregivers’ coping abilities have all been found to contribute to higher levels of family burden.^12, 13, 14^ Lower independence performance, lower self-control attributed to the patients, decrease in social interests, and less affective support, predict burden.^12, 13, 14^

Research indicates that a higher number of patient needs is linked to increased family burden in areas such as daily activities, disrupted behavior, and impact on caregiver routines.^12, 13^ Patient needs that contribute most to family burden include daytime activities, medication, self-care, alcohol, psychotic symptoms, financial issues, and household management.^9^ Other predictors of higher family burden levels include higher levels of psychopathology and disability, as well as being male and older.^12^

Understanding the factors that contribute to caregiver burden is crucial for developing effective interventions and support strategies to improve their quality of life. Therefore, there is a critical need for research to identify the factors that contribute to the caregiving burden in schizophrenia, particularly in Bangladesh. Such knowledge can help healthcare professionals in developing effective interventions to support and empower families in their caregiving role and ultimately reduce the burden on caregivers. This study aims to address this research gap by examining the factors associated with the caregiving burden among family caregivers of individuals with schizophrenia in Bangladesh. The findings of this study will be valuable in guiding the development of targeted interventions that can improve the well-being of both patients and their families.

## Methods

### Study Design

The study was a descriptive cross-sectional study conducted from January to December 2020 at the National Institute of Mental Health and Dhaka Medical College and Hospital in Dhaka, Bangladesh. The study aimed to assess the burden of primary caregivers and associated factors in caring for patients with schizophrenia. The study population included adult caregivers of diagnosed schizophrenia patients who provided written informed consent and were free from any mental illness. Participants were selected using purposive non-probability sampling based on inclusion and exclusion criteria.

### Sample Size Calculation

The desired level of precision was set at 0.05. The prevalence of burden of caregivers was assumed to be 70% based on a previous study.^14^ The reliability coefficient at a 95% confidence interval was set at 1.96. Using the formula n=(z^(2) pq)/d^2, the calculated sample size was 323. However, due to the COVID-19 pandemic, data was collected from 175 respondents.

### Data Collection Procedure

Data was collected through a semi-structured questionnaire and checklist, covering personal information, socio-demographics, and schizophrenia-related details. The validated Bangla version of the Zarit Interview Burden scale assessed caregivers’ burden (with scores ranging from 0 to 88), while a checklist captured illness duration and severity. Zarit Burden Interview is a tool used to measure the burden of caregiving experienced by primary caregivers. The interview consists of 22 items, each rated on a 5-point scale ranging from 0 (Never) to 4 (Nearly always).The cumulative score can range from 0 to 88, with higher scores indicating greater caregiver burden. The interpretation of the ZBI score is as follows: scores of 0–21 indicate little or no burden, 21–40 indicate mild-to-moderate burden, 41–60 indicate moderate-to-severe burden, and scores of 61–88 indicate severe burden. The caregiver is asked to endorse each statement based on their experience of providing care. Interviews were face-to-face by the researcher at the study place, with consent and confidentiality ensured. A pre-test was conducted before data collection at different study sites.

### Pre-testing

Pre-testing was conducted on the questionnaire with 15 primary caregivers of schizophrenic patients and medical reports were also reviewed to ensure its validity. Based on the findings, necessary modifications were made to the questionnaire before data collection at the study places.

### Data Quality Control and Assurance

The study protocol was developed following the standard format of the Institutional Review Board (IRB) of NIPSOM and an extensive literature review was conducted to ensure its quality. Data collection instruments were pre-tested and corrected as necessary, with a validated Bengali questionnaire used for data collection. To ensure data quality, collected data were checked, cleaned, edited, compiled, coded, and categorized according to objectives and variables. Errors were detected and corrected, and the data were entered into a computer for analysis.

### Data Analysis

The collected data were analyzed using SPSS version 25. Descriptive analysis was conducted to determine the frequency, percentage, mean, and standard deviation of the variables. Inferential analysis, such as chi-square test and logistic regression analysis, were used to assess the association between caregiver burden and associated factors. Statistical significance was set at p<0.05.

### Ethical Considerations

The study adhered to the ethical principles outlined in the Declaration of Helsinki and was approved by the National Institute of Preventive and Social Medicine (NIPSOM). Participants provided informed written consent prior to data collection, and their confidentiality and privacy were ensured throughout the study. Participants were also informed of their right to withdraw from the study at any time without penalty.

## Results

The study included a total of 175 adult caregivers of individuals diagnosed with schizophrenia. Sociodemographic information of both caregivers and patients were collected.

### Sociodemographic Characteristics of Caregivers

The study collected socio-demographic information from the 175 caregivers. Most of the caregivers were between 26 to 41 years old (52.6%), female (71.4%), and married (66.9%). The majority had completed primary education (30.9%) and were housewives (49.1%). Their monthly family income was mostly between 5000-15000 taka (69.1%), and 57.1% were from rural areas. Almost half (48.6%) lived in a semi-pakka house, and 50.9% belonged to a joint family. The majority of the caregivers were spouses (28.6%) or parents (24.6%) of the patients. [Table 1]

**Table 1:** Sociodemographic Characteristics of Caregivers:

| **Socio-Demographic Characteristics of Caregivers** | **Percentage** |
| --- | --- |
| **Age** (Mean ±SD: 34.02±10.45 years) | |
| 18-25 years | 25.1% |
| 26-33 years | 25.7% |
| 34-41 years | 26.9% |
| 42-49 years | 9.7% |
| 50 and above years | 12.6% |
| **Gender** |  |
| Female | 71.4% |
| Male | 28.6% |
| **Marital Status** |  |
| Married | 66.9% |
| Unmarried | 23.4% |
| Widow/Widower | 9.7% |
| **Education Status** |  |
| Uneducated | 11.4% |
| Primary Education | 30.9% |
| SSC | 18.9% |
| HSC | 16.6% |
| Graduation | 13.1% |
| Post-Graduation | 9.1% |
| **Occupation** |  |
| Housewives | 49.1% |
| Businessmen | 17.1% |
| Private Job Holders | 12% |
| Students | 11.4% |
| Farmers | 2.9% |
| Physicians | 2.3% |
| Government Service Holders | 2.3% |
| Unemployed | 2.3% |
| Retired | 0.6% |
| **Monthly Family Income** |  |
| <5000 Taka | - |
| 5000-15000 Taka | Majority |
| 15000-25000 Taka | 16.6% |
| >25000 Taka | 14.3% |
| **Place of Residence** |  |
| Urban | 42.9% |
| Rural | 57.1% |
| **Condition of House** |  |
| Kacha (clay house) | 20.6% |
| Semi-pakka (semi-solid) | 48.6% |
| Pakka (solid) | 30.9% |
| **Type of Family** |  |
| Joint Family | 50.9% |
| Single Family | 49.1% |
| **Number of Family Members** |  |
| 2-5 Members | 48.3% |
| 6-9 Members | 30.3% |
| 10+ Members | 11.4% |
| **Relationship with Patients** |  |
| Spouses | 28.6% |
| Parents | 24.6% |
| Children | 20% |
| Siblings | 6.9% |
| Others | 20% |

### Sociodemographic Characteristics of Patients

The majority (28%) of patients were aged between 34 to 41 years, and the mean age was 40.19±10.43 years. In terms of gender, 56% of patients were male and 44% were female. [Table 2]

**Table 2:** Sociodemographic Characteristics and Disease Condition Information of Patients Cared for by Respondents.

| **Sociodemographic Characteristics of Patients** |  |
| --- | --- |
| **Variables** | **Percentage** |
| **Age** (Mean ±SD: $40.19 \pm 10.43$) | |
| 18-25 years | 10.5% |
| 26-33 years | 23.5% |
| 34-41 years | 28% |
| 42-49 years | 16.5% |
| 50 and above | 21.5% |
| **Gender** |  |
| Female | 44% % |
| Male | 56% % |
| **Disease condition information of the patients cared for by respondents** |  |
| **Stage of illness** |  |
| Stage 1 | 57.7% |
| Stage 2 | 36% |
| Stage 3 | 6.3% |
| **Duration of illness (Mean ± SD: 4.10±3.26)** |  |
| <5 years | 66.3% |
| ≥5 years | 33.7% |

### Disease condition information of the patients cared for by respondents

Stage and duration of illness of the patients cared for by respondents:

The stage of illness of the patients cared for by the respondents was categorized as stage 1, stage 2, and stage 3. The majority of patients (57.7%) had stage 1 illness, while 36% had stage 2 and 6.3% had stage 3 illness. The majority of patients (66.3%) had an illness duration of less than 5 years, while 33.7% had an illness duration of 5 years or more. The mean duration of illness was 4.10±3.26 years. [Table 2]

### Burden Profile of Caregivers Assessed Using the Zarit Burden Interview (ZBI)

Caregiver burden was analyzed based on their ZBI scores. Out of 175 respondents, only 1.7% reported little or no burden, 21.7% reported mild-to-moderate burden, 62.3% reported moderate-to-severe burden, and 14.3% reported severe burden. The mean ZBI score was 49.49±12.06, indicating a moderate-to-severe burden among the majority of caregivers. [Table 3]

**Table 3:** Caregiver Burden as Assessed by Zarit Burden Interview Scores.

| **ZBI Score Range** | **Percentage (%)** |
| --- | --- |
| 0-21 (Little or no burden) | 1.7 |
| 21-40 (Mild-to-moderate burden) | 21.7 |
| 41-60 (Moderate-to-severe burden) | 62.3 |
| 61-88 (Severe burden) | 14.3 |
| Mean±SD | 49.49±12.06 |

### Relationship between Caregiver Burden and Sociodemographic Characteristics

The study analyzed the association between the burden of respondents and different sociodemographic variables. The sociodemographic variables were categorized based on the level of burden. Age (p=0.05), gender (p=0.01), occupation (p=0.01), monthly income (p=0.01), marital status (p=0.01), condition of the house (p=0.01), and relationship with patients were associated with caregiver burden (p=0.01). Respondents aged 26 to 33 years had the highest mild to moderate burden (44.7%), while those aged 34 to 41 years had the highest moderate to severe burden (27.6%). Female respondents had a higher moderate to severe burden than male respondents (76.1% vs 23.9%). Housewives had the highest moderate to severe burden (54.1%). Respondents with a monthly family income of 5000-15000 taka had the highest moderate to severe burden (72.5%). Married respondents had the highest mild to moderate burden (69.7%). Respondents living in a semi-pakka house had the highest moderate to severe burden (51.4%). Respondents who had a spouse relationship with a mentally ill patient had the highest moderate to severe burden (32.1%). [Table 4] The association between mean burden score and sociodemographic factors are also shown in table 4.

**Table 4:**
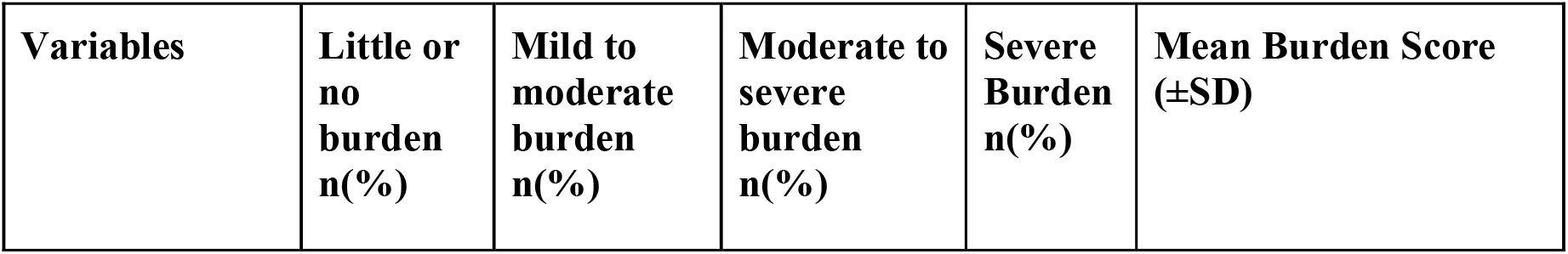
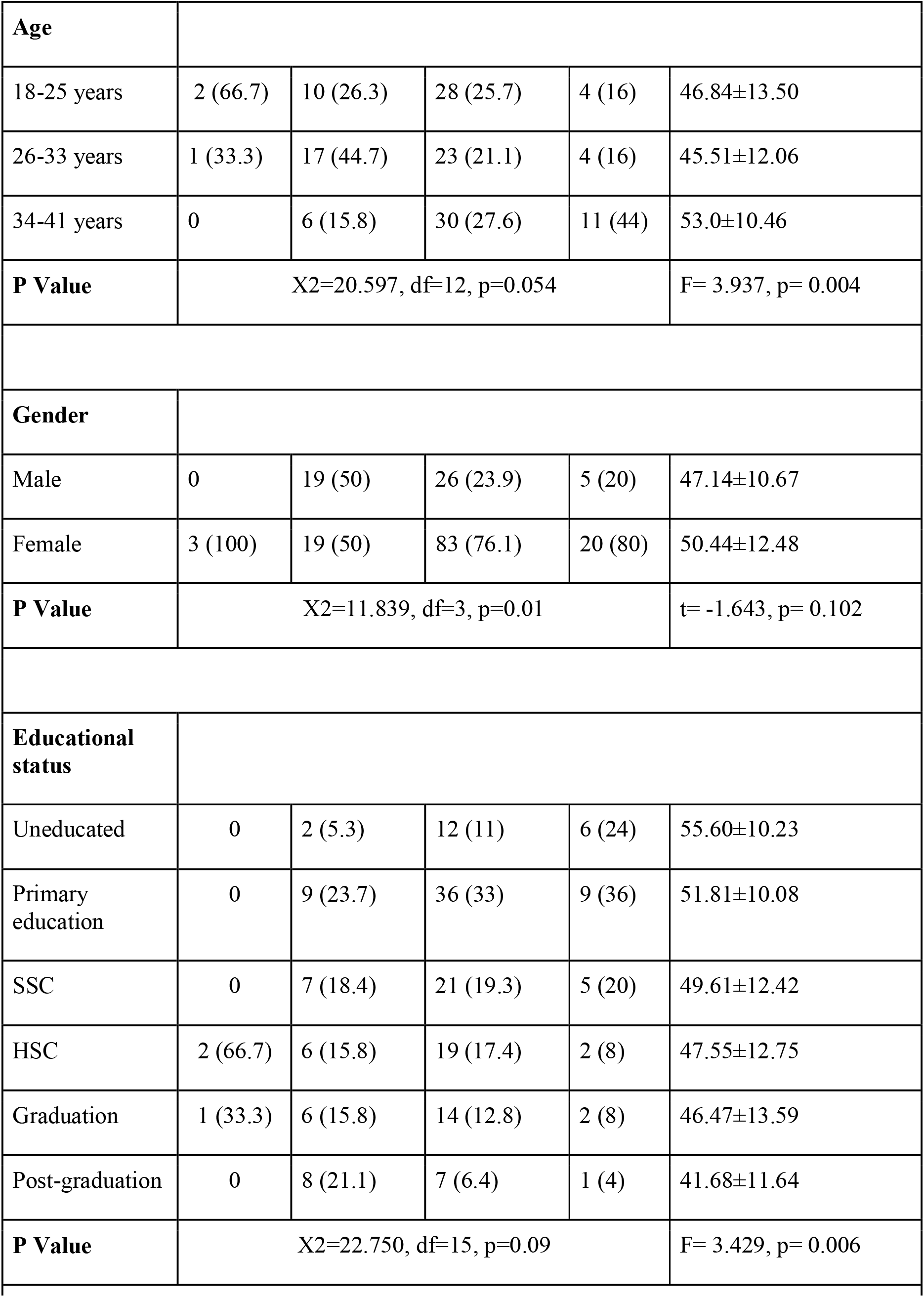
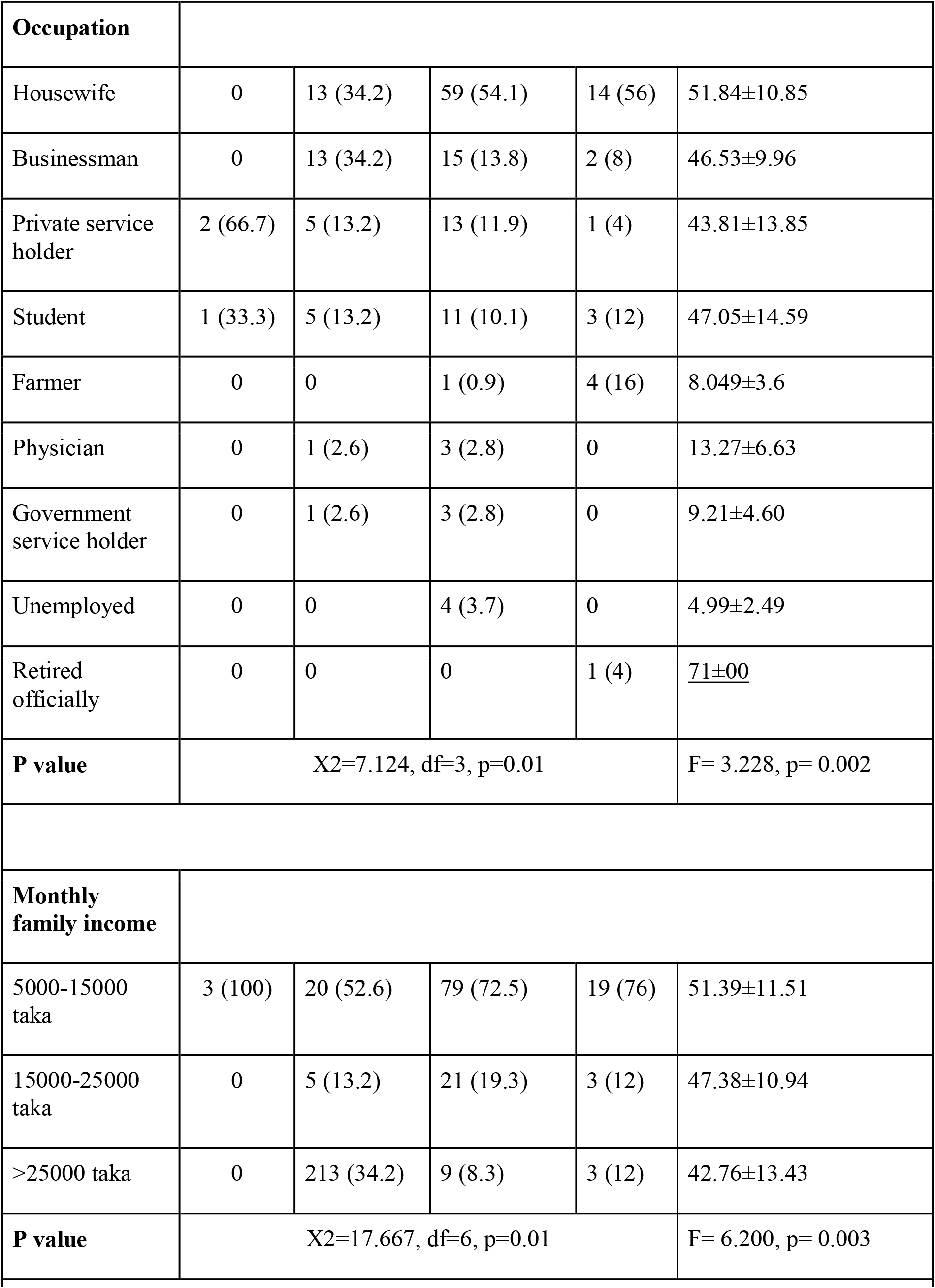
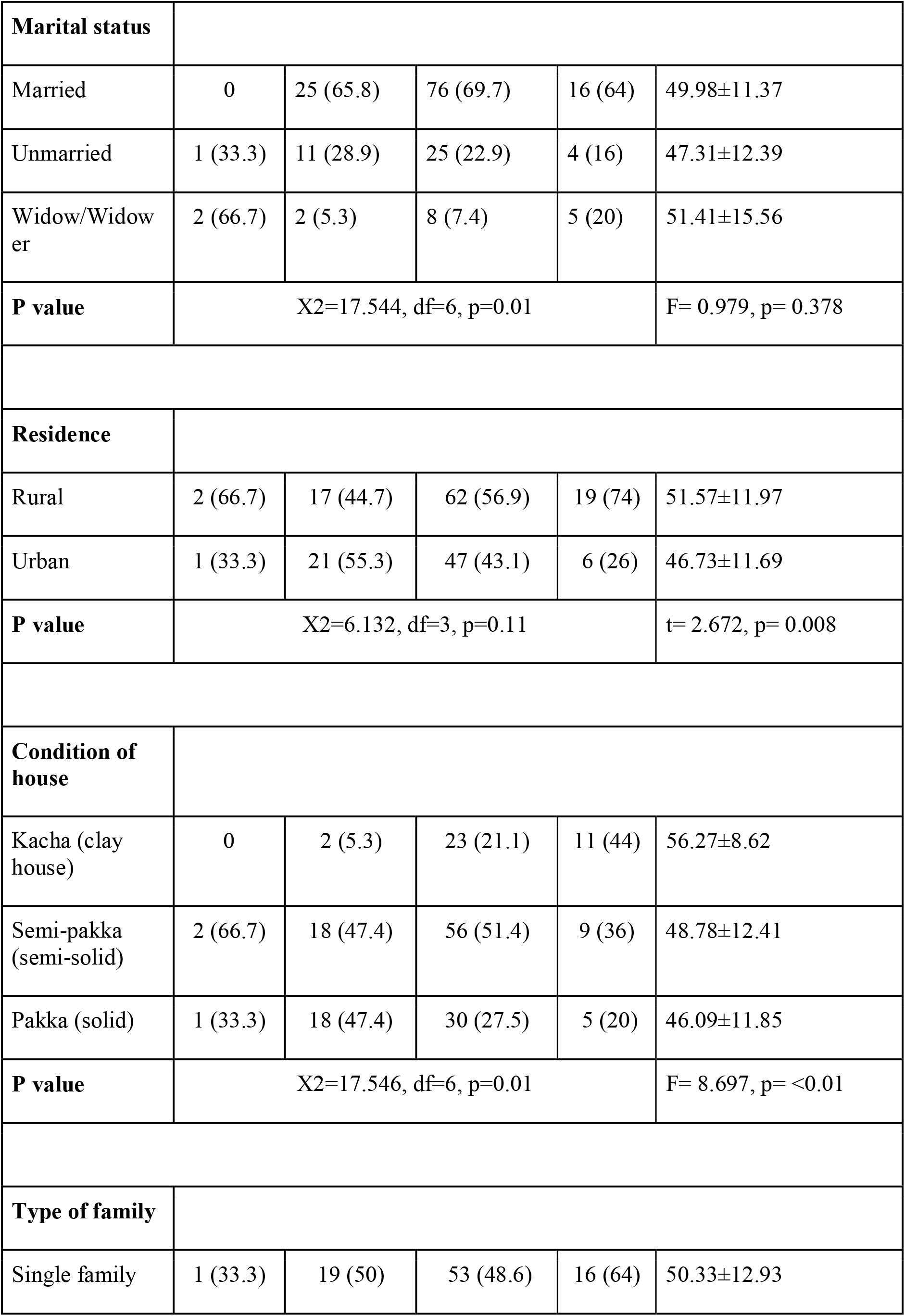
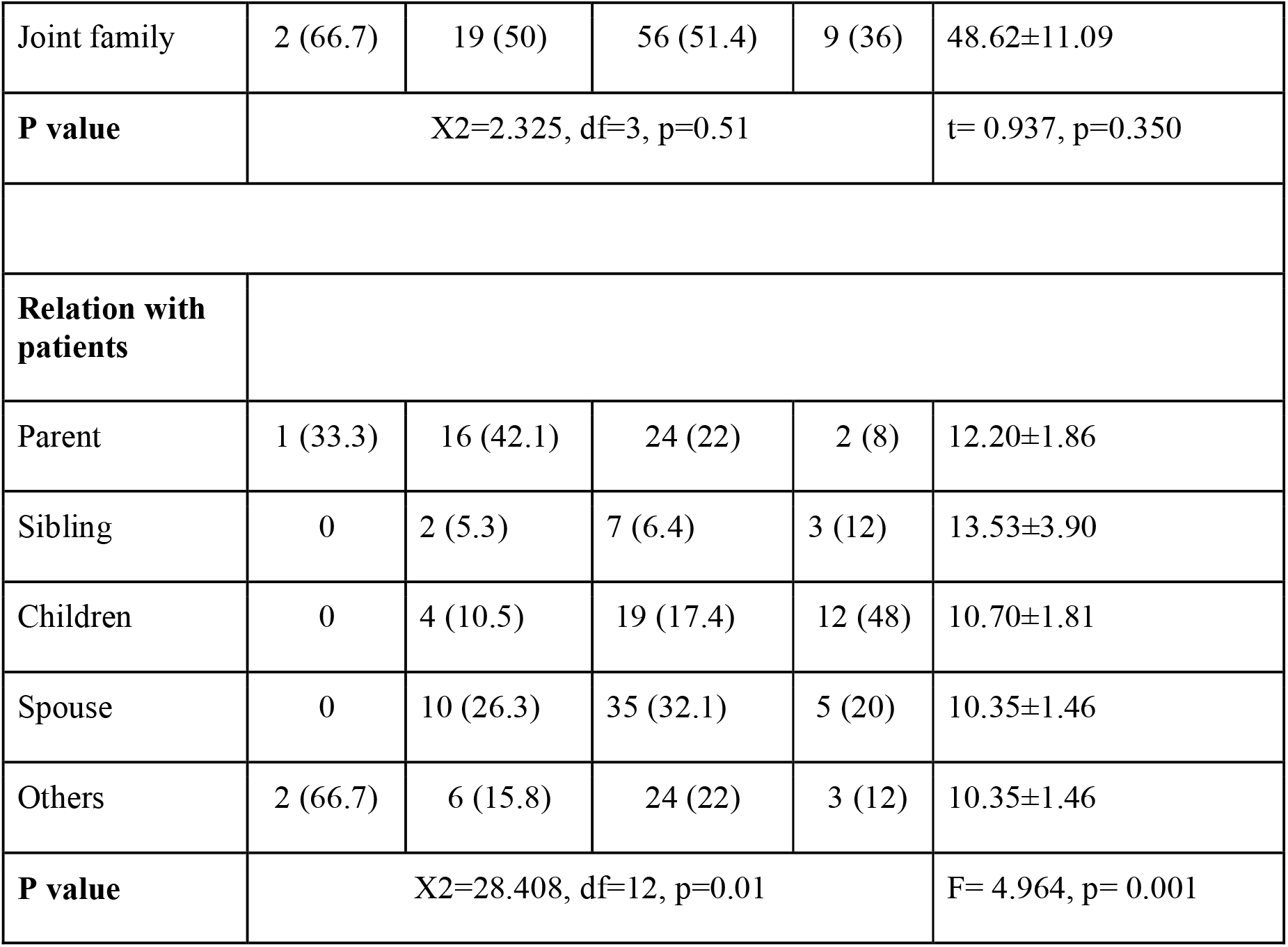
Association of caregiver burden with sociodemographic variables of caregivers.

| **Variables** | **Little or no burden**<br>**n(%)** | **Mild to moderate burden**<br>**n(%)** | **Moderate to severe burden**<br>**n(%)** | **Severe Burden**<br>**n(%)** | **Mean Burden Score**<br>**(±SD)** |
| --- | --- | --- | --- | --- | --- |
| Age |  |  |  |  |  |
| 18-25 years | 2 (66.7) | 10 (26.3) | 28 (25.7) | 4 (16) | 46.84±13.50 |
| 26-33 years | 1 (33.3) | 17 (44.7) | 23 (21.1) | 4 (16) | 45.51±12.06 |
| 34-41 years | 0 | 6 (15.8) | 30 (27.6) | 11 (44) | 53.0±10.46 |
| P Value | X2=20.597, df=12, p=0.054 |  |  |  | F= 3.937, p= 0.004 |
| Gender |  |  |  |  |  |
| Male | 0 | 19 (50) | 26 (23.9) | 5 (20) | 47.14±10.67 |
| Female | 3 (100) | 19 (50) | 83 (76.1) | 20 (80) | 50.44±12.48 |
| P Value | X2=11.839, df=3, p=0.01 |  |  |  | t= -1.643, p= 0.102 |
| Educational status |  |  |  |  |  |
| Uneducated | 0 | 2 (5.3) | 12 (11) | 6 (24) | 55.60±10.23 |
| Primary education | 0 | 9 (23.7) | 36 (33) | 9 (36) | 51.81±10.08 |
| SSC | 0 | 7 (18.4) | 21 (19.3) | 5 (20) | 49.61±12.42 |
| HSC | 2 (66.7) | 6 (15.8) | 19 (17.4) | 2 (8) | 47.55±12.75 |
| Graduation | 1 (33.3) | 6 (15.8) | 14 (12.8) | 2 (8) | 46.47±13.59 |
| Post-graduation | 0 | 8 (21.1) | 7 (6.4) | 1 (4) | 41.68±11.64 |
| P Value | X2=22.750, df=15, p=0.09 |  |  |  | F= 3.429, p= 0.006 |

| Occupation |  |  |  |  |  |
| --- | --- | --- | --- | --- | --- |
| Housewife | 0 | 13 (34.2) | 59 (54.1) | 14 (56) | 51.84±10.85 |
| Businessman | 0 | 13 (34.2) | 15 (13.8) | 2 (8) | 46.53±9.96 |
| Private service holder | 2 (66.7) | 5 (13.2) | 13 (11.9) | 1 (4) | 43.81±13.85 |
| Student | 1 (33.3) | 5 (13.2) | 11 (10.1) | 3 (12) | 47.05±14.59 |
| Farmer | 0 | 0 | 1 (0.9) | 4 (16) | 8.049±3.6 |
| Physician | 0 | 1 (2.6) | 3 (2.8) | 0 | 13.27±6.63 |
| Government service holder | 0 | 1 (2.6) | 3 (2.8) | 0 | 9.21±4.60 |
| Unemployed | 0 | 0 | 4 (3.7) | 0 | 4.99±2.49 |
| Retired officially | 0 | 0 | 0 | 1 (4) | <u>71±00</u> |
| **P value** | X2=7.124, df=3, p=0.01 |  |  |  | F= 3.228, p= 0.002 |
| Monthly family income |  |  |  |  |  |
| 5000-15000 taka | 3 (100) | 20 (52.6) | 79 (72.5) | 19 (76) | 51.39±11.51 |
| 15000-25000 taka | 0 | 5 (13.2) | 21 (19.3) | 3 (12) | 47.38±10.94 |
| >25000 taka | 0 | 213 (34.2) | 9 (8.3) | 3 (12) | 42.76±13.43 |
| **P value** | X2=17.667, df=6, p=0.01 |  |  |  | F= 6.200, p= 0.003 |
| Marital status |  |  |  |  |  |
| Married | 0 | 25 (65.8) | 76 (69.7) | 16 (64) | 49.98±11.37 |
| Unmarried | 1 (33.3) | 11 (28.9) | 25 (22.9) | 4 (16) | 47.31±12.39 |
| Widow/Widow er | 2 (66.7) | 2 (5.3) | 8 (7.4) | 5 (20) | 51.41±15.56 |
| P value | X2=17.544, df=6, p=0.01 |  |  |  | F= 0.979, p= 0.378 |
| Residence |  |  |  |  |  |
| Rural | 2 (66.7) | 17 (44.7) | 62 (56.9) | 19 (74) | 51.57±11.97 |
| Urban | 1 (33.3) | 21 (55.3) | 47 (43.1) | 6 (26) | 46.73±11.69 |
| P value | X2=6.132, df=3, p=0.11 |  |  |  | t= 2.672, p= 0.008 |
| Condition of house |  |  |  |  |  |
| Kacha (clay house) | 0 | 2 (5.3) | 23 (21.1) | 11 (44) | 56.27±8.62 |
| Semi-pakka (semi-solid) | 2 (66.7) | 18 (47.4) | 56 (51.4) | 9 (36) | 48.78±12.41 |
| Pakka (solid) | 1 (33.3) | 18 (47.4) | 30 (27.5) | 5 (20) | 46.09±11.85 |
| P value | X2=17.546, df=6, p=0.01 |  |  |  | F= 8.697, p= <0.01 |
| Type of family |  |  |  |  |  |
| Single family | 1 (33.3) | 19 (50) | 53 (48.6) | 16 (64) | 50.33±12.93 |
| Joint family | 2 (66.7) | 19 (50) | 56 (51.4) | 9 (36) | 48.62±11.09 |
| **P value** | X2=2.325, df=3, p=0.51 |  |  |  | t= 0.937, p=0.350 |
| **Relation with patients** |  |  |  |  |  |
| Parent | 1 (33.3) | 16 (42.1) | 24 (22) | 2 (8) | 12.20±1.86 |
| Sibling | 0 | 2 (5.3) | 7 (6.4) | 3 (12) | 13.53±3.90 |
| Children | 0 | 4 (10.5) | 19 (17.4) | 12 (48) | 10.70±1.81 |
| Spouse | 0 | 10 (26.3) | 35 (32.1) | 5 (20) | 10.35±1.46 |
| Others | 2 (66.7) | 6 (15.8) | 24 (22) | 3 (12) | 10.35±1.46 |
| **P value** | X2=28.408, df=12, p=0.01 |  |  |  | F= 4.964, p= 0.001 |

### Relationship between Caregiver Burden and Patient Disease Conditions, Based on Burden Level and Average Burden Score

The burden score of caregivers was significantly associated with the stage and duration of illness of the patients. Caregivers of patients with stage 1 illness had the highest moderate to severe burden (72.5%). Caregivers of patients with an illness duration of below than 5 years had the highest moderate to severe burden (66.1%). Caregivers of patients with an illness duration of more than 5 years had higher burden scores compared to those with a duration of 5 years or less. [Table 5]

**Table 5:** Association of caregiver burden with disease conditions of patients.

| Disease condition | Little or no burden n | Mild to moderate | Moderate to severe | Severe Burden | Mean Burden Score (±SD) |
| --- | --- | --- | --- | --- | --- |

|  | (%) | burden n (%) | burden n (%) | (%) |  |
| --- | --- | --- | --- | --- | --- |
| Stage of Illness |  |  |  |  |  |
| Stage 1 | 3 (100) | 31 (81.6) | 58 (53.2) | 9 (36) | 12.51±1.24 |
| Stage 2 | 0 | 7 (18.4) | 44 (40.4) | 12 (48) | 9.83±1.23 |
| Stage 3 | 0 | 0 | 7 (6.4) | 4 (16) | 5.87±1.77 |
| P Value | X2=19.358, df=6, p=0.01 |  |  |  | F= 12.918 p= <0.01 |
| Duration of Illness |  |  |  |  |  |
| ≤5 years | 3 (100) | 31 (81.6) | 72 (66.1) | 10 (40) | 47.43±12.35 |
| >5 years | 0 | 7 (18.4) | 37 (33.9) | 15 (60) | 53.54±10.42 |
| P Value | X2=13.235, df=3, p=0.01 |  |  |  | t= -3.251, p= 0.001 |

## Discussion

The current study aimed to explore the sociodemographic characteristics of caregivers of individuals diagnosed with schizophrenia and their relationship with caregiver burden. The study included 175 adult caregivers who provided care for individuals diagnosed with schizophrenia. The majority of caregivers were female, married, and had completed primary education. Additionally, the majority of patients were male and aged between 34 to 41 years, with stage 1 illness being the most common. The study found that the majority of caregivers experienced moderate-to-severe burden, as indicated by their ZBI scores. The findings of this study align with previous research indicating that caregivers of individuals with schizophrenia often experience significant burden. Existing literature indicates that caregivers of individuals with schizophrenia experience high levels of subjective burden.^15^ Moreover, research has found that the severity of positive psychotic symptoms is associated with significant objective burden, including financial strain, increased family conflicts, and mental health difficulties and reduced life satisfaction.^16^

Furthermore, the study investigated the relationship between caregiver burden and sociodemographic characteristics of the caregivers. The results indicated that sociodemographic variables, such as age, gender, occupation, monthly income, marital status, and condition of the house, were associated with caregiver burden. Female respondents, housewives, and those with lower monthly family income experienced higher burden scores. Additionally, the burden score of caregivers was significantly associated with the stage and duration of illness of the patients. The current study found that the majority of caregivers of individuals diagnosed with schizophrenia experienced moderate-to-severe burden, as indicated by their ZBI scores. The mean ZBI score was 49.49±12.06, with 62.3% reporting moderate-to-severe burden, 21.7% reporting mild-to-moderate burden, and only 1.7% reporting little or no burden. Caregivers of patients with stage 1 illness and those with an illness duration of less than 5 years experienced higher burden scores. Other studies using the ZBI instrument have reported varying levels of caregiver burden, with some reporting higher levels of severe burden^14,17^, while others reported varying levels.^18,19^ A potential explanation for the higher ZBI score observed in this study compared to others may be the limited availability of facilities for caring for patients in Bangladesh. This could potentially increase the burden on caregivers and contribute to the higher scores observed.

The study found that 71.4% of the caregivers were female. This finding is consistent with another study by Shamsaei et al., which also reported a predominance of female caregivers at 73.7%.^17^ In the current study, over half (57.1%) of the caregivers were from rural areas, with a majority being married (66.9%), and most were housewives (49.1%) or employed (36.6%). The majority (50.9%) lived in joint families and semi-pakka houses (48.6%), with 58.3% having 2-5 family members. The previous study by Bhat et al. also found that a majority of caregivers were from rural areas (64%) and married (84%), with low educational backgrounds and most having a family size between 5 and 10 (76%).^14^ Other studies by Shamsaei et al., and Souza et al. also reported similar findings.^17, 19^

The majority (66.3%) of patients had an illness duration of <5 years, while 33.7% had an illness duration of ≥5 years, with a mean duration of illness of 4.10±3.26 years. Most patients (57.7%) had Stage 1 illness, with 36% having Stage 2 and 6.3% having Stage 3 illness. However, these results may differ from other studies due to differences in enrollment during the study period. For instance, Bhat et al. found that approximately 70% of patients had an illness duration of 5 years or more.^14^ In this study, the stage and duration of illness were significantly associated with caregiver burden. This finding was similar to that of Shamsaei et al., who found that years of chronic illness and caregiving duration were significant factors that influenced caregiver burden.^17^ In contrast, Zarit et al. found that illness duration was not related to burden.^20^ The factors that increased caregiver burden in this study were similar to some previous studies, while different from others. These differences could be attributed to differences in methodology, cultural differences in these countries, and sociodemographic characteristics of caregivers.^19^

These findings highlight the need to provide support to caregivers of individuals diagnosed with schizophrenia, particularly those who are female, housewives, and have lower income. Furthermore, healthcare professionals should consider the stage and duration of illness when providing support to caregivers, as these factors have a significant impact on caregiver burden. Overall, the study provides important insights into the factors that contribute to caregiver burden among caregivers of individuals diagnosed with schizophrenia, and may help inform interventions aimed at reducing this burden.

## Conclusion

The study highlights the burden experienced by primary caregivers of schizophrenia patients and its associated factors. Community interventions such as redesigning in-home services are necessary to meet the social needs of patients and provide respite to caregivers. Health systems should conduct a mapping of psychosocial provisions to decrease family burden rates and improve transition to society. Future research could involve a larger sample size and a comparative study between rural and urban areas. Overall, this study provides insights into the burden experienced by caregivers of schizophrenia patients and can inform interventions to address this issue.

## Data Availability

All data produced in the present study are available upon reasonable request to the authors.

## Abbreviations

ZBI: Zarit Burden Interview
IRB: Institutional Review Board
NIPSOM: National Institute of Preventive and Social Medicine
SPSS: Statistical Package for Social Sciences

## Acknowledgments

Not applicable.

## Authorship Contribution

Conceptualization: TTT, NR, SSH, VP; Data curation:TTR, NR, SR; Formal analysis: TTT; Investigation: TTT, NR, SSH; Methodology: TTT, NR, MG; Project administration: TTT, NR, SSH; Software: TTT, NR; FA; Supervision: TTT, NR, SSH; Validation: TTT; Visualization; Roles/Writing - original draft: TTT, VP, NR, SSH, FAL, SPK; Writing - review & editing: TTT, NR, VP, SSH

## ICMJE Statement

This article complies with the International Committee of Medical Journal Editors’ uniform requirements for the manuscript.

## Statement of Ethics

Prior to the commencement of this study, ethical approval was obtained from the Institutional Review Board (IRB) of National Institute of Preventive and Social Medicine (NIPSOM). This approval ensured that the study was conducted in an ethical manner, in accordance with relevant guidelines and regulations.

